# The effectiveness of interventions to reduce COVID-19 transmission in a large urban jail

**DOI:** 10.1101/2020.06.16.20133280

**Authors:** Giovanni S. P. Malloy, Lisa Puglisi, Margaret L. Brandeau, Tyler D. Harvey, Emily A. Wang

**Affiliations:** Department of Management Science and Engineering, School of Engineering, Stanford University, Stanford, CA, USA; Department of Internal Medicine, School of Medicine, Yale University, New Haven, CT, USA; Pain Research, Informatics, Multimorbidities and Education Center, VA Connecticut Healthcare System, West Haven, CT, USA

## Abstract

**Objectives:** To estimate the impact of various mitigation strategies on COVID-19 transmission in a U.S. jail beyond those offered in national guidelines.

**Methods:** We developed a stochastic dynamic transmission model of COVID-19 in one large urban U.S. jail among staff and incarcerated individuals. We divided the outbreak into four intervention phases: the start of the outbreak, depopulation of the jail, increased proportion of people in single cells, and asymptomatic testing. We used the next generation method to estimate the basic reproduction ratio, *R*_0_, in each phase. We estimated the fraction of new cases, hospitalizations, and deaths averted by these interventions along with the standard measures of sanitization, masking, and social distancing interventions.

**Results:** For the first outbreak phase, the estimated *R*_0_ was 8.23 (95% CrI: 5.01-12.90), and for the subsequent phases, *R*_0,*phase* 2_ = 3.58 (95% CrI: 2.46-5.08), *R*_0,*phase* 3_ = 1.72 (95% CrI: 1.41-2.12), and *R*_0,*phase* 4_ = 0.45 (95% CrI: 0.32-0.59). In total, the jail’s interventions prevented approximately 83% of projected cases and hospitalizations and 89% of deaths over 83 days.

**Conclusions:** Depopulation, single celling, and asymptomatic testing within jails can be effective strategies to mitigate COVID-19 transmission in addition to standard public health measures.

**Policy Implications:** Decision-makers should prioritize reductions in the jail population, single celling, and testing asymptomatic populations, as additional measures to manage COVID-19 within correctional settings.

## INTRODUCTION

COVID-19, the disease caused by the SARS-CoV-2 virus, has affected millions of people worldwide, with disproportionate impact on some communities such as those inside correctional facilities. In the United States (U.S.), approximately 2.2 million people are incarcerated in any given day in over 5,000 facilities,^1^ where the built environment and activities of daily living make physical distancing exceedingly difficult to implement.^2,3^ As of the third week of April 2020, 420 U.S. correctional facilities had at least one diagnosed case of COVID-19, accounting for a total of 4,893 cases among incarcerated individuals and 2,778 cases among staff members.^3^ As of June, correctional facilities account for eight of out ten of the largest COVID-19 outbreaks nationally, surpassing nursing homes and food processing plants, and 26 states now have a higher rate of COVID-19 infection in their correctional population than in their general population.^4,5^ Cook County Jail currently has one of the largest outbreak in the country, and the infection rate at Rikers Island is nearly five times that of New York City.^6,7^

Despite the severity of outbreaks in correctional facilities, national guidance surrounding the prevention and management of COVID-19 within such settings has been limited. In the weeks after the first major outbreak in a U.S. jail, the Centers for Disease Control and Prevention (CDC) published policy guidelines for correctional facilities to help mitigate COVID-19 transmission, which included limiting transfer of incarcerated people between facilities, restricting the number of visitors entering facilities, promoting personal hygiene and environmental sanitization, maximizing the space between those incarcerated (i.e. arranging bunks so individuals sleep head to toe), and screening staff for symptoms.^8^

However, CDC guidelines then and still now do not account for the difficulty that many facilities face in managing COVID-19 and creating physical distance within jails. Even among those jails which are not crowded, physical distancing is challenging given use of congregate living arrangements, shared meals, and exercise and recreation programming. In the absence of more targeted guidelines, there is wide variance in how correctional facilities are managing COVID-19, especially regarding depopulation efforts that may mitigate COVID-19 and approaches to testing (symptomatic only vs. asymptomatic, viral testing vs. antibody testing). As an example, Attorney General Barr has ordered that medically frail individuals in federal prisons be released to home quarantine, whereas many state prison systems have no stated policies for larger scale release. Some correctional systems have implemented systemwide testing of all incarcerated individuals, including those who are asymptomatic, while others are only testing those who are symptomatic.

The effectiveness of such measures, which fall outside of CDC guidance, in reducing the transmission of COVID-19 within correctional facilities has yet to be established. In this study, we estimate the effectiveness of measures to mitigate the spread of COVID-19 beyond standard CDC recommendations in a large urban jail. We focus on policies with large potential impact for which there is variability in practice, namely depopulation (cessation of new detentions and release of incarcerated individuals), single celling (percentage of the total incarcerated population in a single cell), and testing asymptomatic individuals with the aim of providing guidance to correctional policymakers and public health agencies.

## METHODS

We developed a stochastic dynamic transmission model of COVID-19 which we calibrated to the outbreak in the jail. We combined data on cases in incarcerated people and correctional staff because they interact very closely and regularly as an ecosystem behind the walls of the jail. We divided the outbreak timeline into four intervention phases marked by the start of the outbreak, start of depopulation efforts, increased single celling, and large-scale asymptomatic testing of incarcerated individuals. We estimated the initial basic reproduction ratio, *R*_0_, and the effective reproduction ratio, *R*_*t*_, in each phase, for the entire jail. We also estimated the fraction of new cases, hospitalizations, and deaths averted by the combined interventions.

### Model description

We modified a traditional SEIR model to represent the disease states of COVID-19. These disease states included susceptible (*S*), exposed (*E*), infected symptomatic (*I*_*sym*_), infected asymptomatic (*I*_*asym*_), quarantined (*Q*), hospitalized (*H*), and recovered (*Rec*) individuals (Figure 1). To model these interacting populations, we developed a mass-action mixing model described by the following equations:

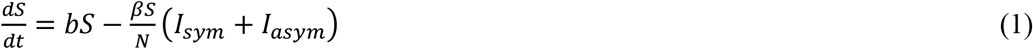

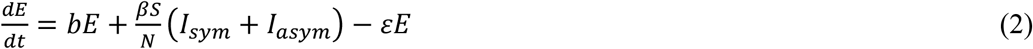

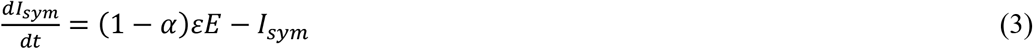

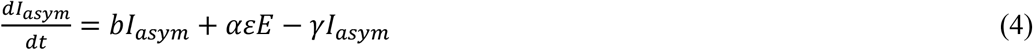

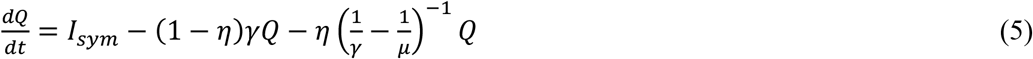

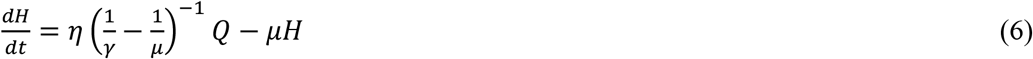

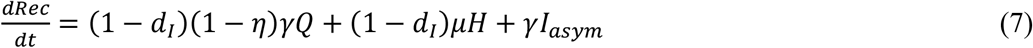

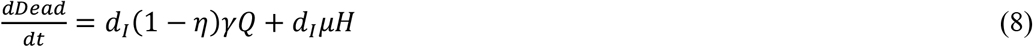

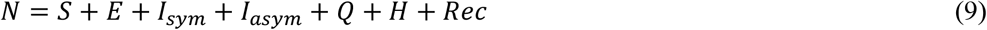

**Figure 1.**
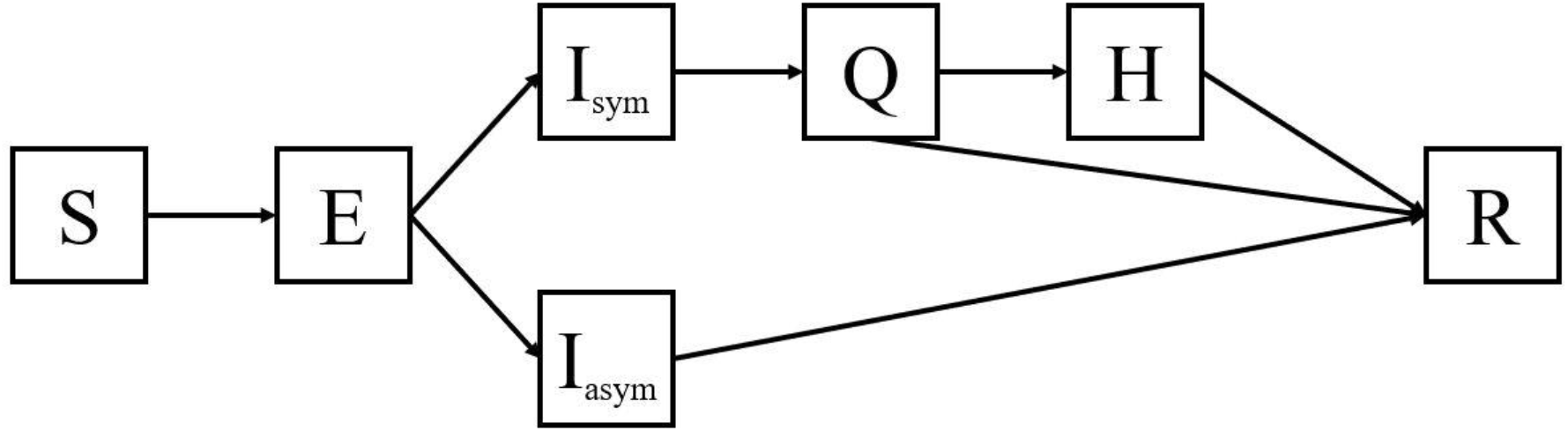
Structure of the disease transmission model.

The susceptible, exposed, and asymptomatic infected populations grew at rate *b* which represented the overall growth or reduction in jail population. We assume that symptomatic infected individuals are not removed from the jail during depopulation and would be admitted directly to quarantine. For the time horizon of the model, the population was generally shrinking. Susceptible individuals were exposed to COVID-19 at transmission rate *β*. We re-calibrated this transmission rate for each of the four outbreak phases. We assumed that asymptomatic and symptomatic infected individuals could transmit the disease.^9,10^ Exposed individuals were infected but not yet infectious and become asymptomatic or symptomatic infected at rate *ε*, which corresponded to the incubation period of COVID-19. A certain proportion, *α*, of these individuals stayed asymptomatic, while remaining individuals became symptomatic. Based on the jail’s report, we assumed that symptomatic infected individuals were placed in quarantine after one day. We assumed that individuals once quarantined did not transmit COVID-19. A fraction, *η*, of quarantined individuals were hospitalized and recovered from hospitalization at rate *μ*. All infected individuals recovered or died at rate *γ* regardless of symptomatic or asymptomatic status. Symptomatic infected individuals died with probability *d*_*I*_.

### Interventions

The jail implemented various measures over time in an attempt to mitigate the spread of COVID-19. We divide the outbreak into four intervention phases, corresponding to the initiation of key measures of interest which fell outside the guidance of the CDC. During Phase 1 (days 1-11), the jail implemented a broad array of strategies that were consistent with CDC guidance including: basic screening for flu-like symptoms in incarcerated people; new detainees quarantined for 14 days and basic screening for flu-like symptoms for visitors, vendors, attorneys, and community members entering the facility; staff required to report symptoms as well as contact with known COVID-19 positive cases and any travel outside of the United States; suspension of all tours, large gatherings, in-person visitation. During phase 2 (days 12-17), the jail population started to decrease by 1.41% each day through a combination of measures which included a marked decrease in new detentions given changes in the court and judicial system procedures and large community organized bail outs (Figure 2). The jail also began taking the temperature of all employees each day. During phase 3 (days 18-36), the jail began increasing the portion of the population in single-occupancy cells from 26% on day 18 to 54% on day 36. During this period, they began requiring all staff to wear surgical masks and allotted new masks to those incarcerated each day. They also continued to isolate confirmed and suspected COVID-19 cases among incarcerated individuals but given the number of individuals, they identified a different building for segregating patients which provided a larger space for the growing number of confirmed cases. Lastly, they started on-site voluntary testing for employees and a two-week COVID-19 paid leave policy for all employees. During phase 4 (days 37-83), the jail began testing for asymptomatic cases in divisions with high numbers of cases identified during contact tracing at a rate of approximately 50-75 people per day.

**Figure 2.**
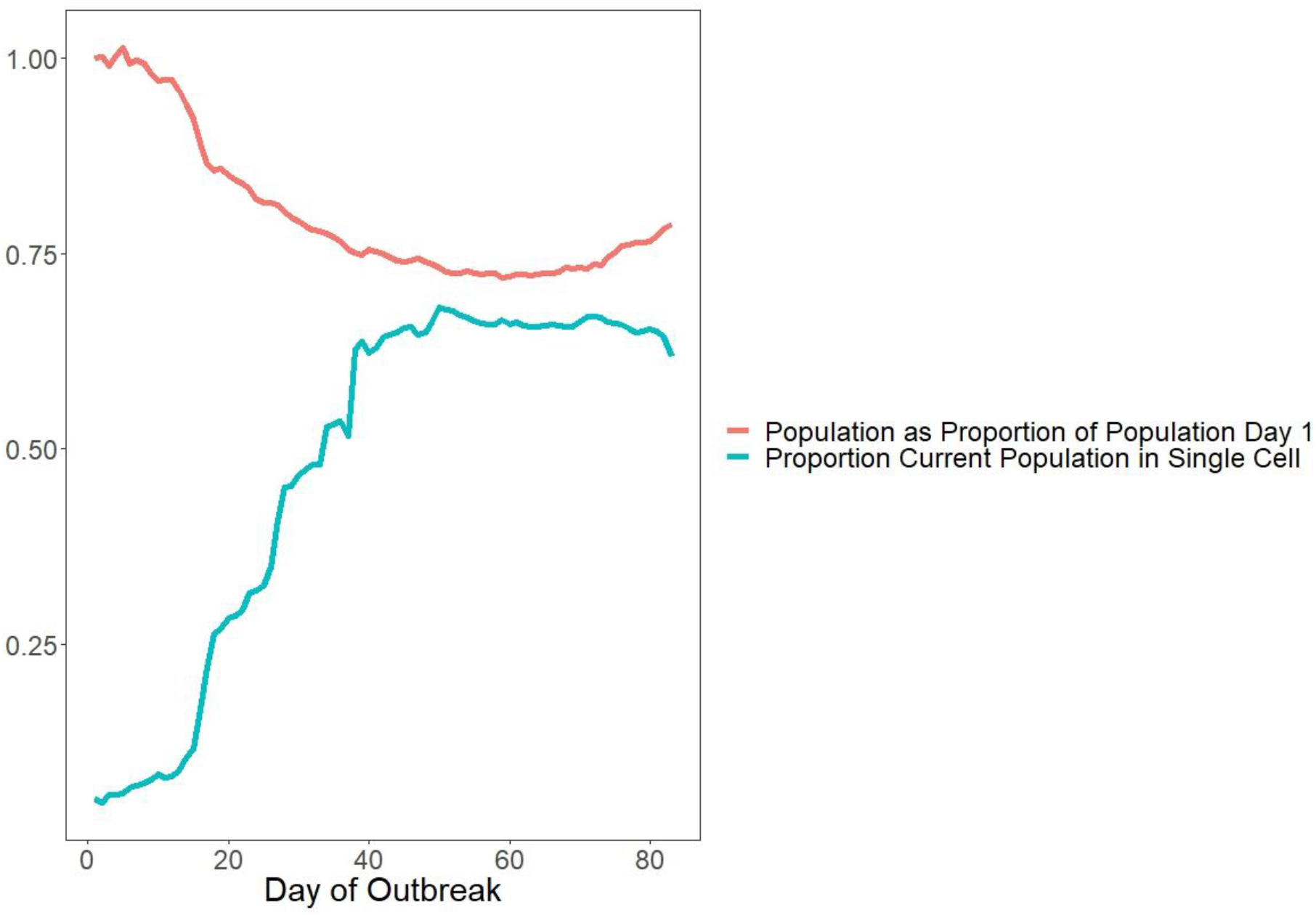
Change in the total population of the jail and the portion of the population in single-occupancy cells over the course of the outbreak.

### Model Instantiation and Calibration

We estimated some model parameter values from previous literature (Table 1). The rate at which exposed individuals became asymptomatically or symptomatically infected, *ε*, was the inverse of the incubation period. The incubation period of COVID-19 was previously described with a lognormal distribution with mean 5.1 days and standard deviation 0.89 days.^11^ We assumed that the proportion of infections that are asymptomatic, *α*, was uniformly distributed over the range 0.25 to 0.56.^12,13^ The average recovery rate was previously estimated to be 0.1, the inverse of the 10-day mean infection period.^14^ We assumed that the infection period followed a truncated normal distribution with mean 10 days, standard deviation 6.25 days, minimum 5 days, and maximum 20 days. Additionally, the length of hospitalization from COVID-19 has been estimated to be 5 days, making the daily recovery probability from the hospital 0.2.^15^ We assumed that the length of hospitalization followed a lognormal distribution with a mean of 5 days and standard deviation of 1 day.

**Table 1.** Parameter Estimates.

| Name | Description | Value | Source |
| --- | --- | --- | --- |
| $b$ | Net rate of entrance into the jail, phase 1 [1/day] | -0.004 | Jail dataset |
|  | Net rate of entrance into the jail, phase 2 [1/day] | -0.0141 |  |
|  | Net rate of entrance into the jail, phase 3 [1/day] | -0.0076 |  |
|  | Net rate of entrance into the jail, phase 4 [1/day] | 0.0005 |  |
| $\beta$ | Transmission rate [1/day] | | Calibrated |
| $\varepsilon$ | Incubation period<sup>-1</sup> [1/day] | 0.18<br>Incubation period: *Lognormal*(5.1, 0.89) | 11 |
| $\alpha$ | Proportion of cases that are asymptomatic | 0.405<br>*uniform*(0.25, 0.56) | 12,13 |
| $\gamma$ | Recovery rate [1/day] | 0.1<br>Infection period:<br>*Truncated N*(10, 6.25, *min* = 5, *max* = 20) | 14,15 |
| $\eta$ | Proportion of symptomatic infections that are hospitalized | 0.14 | Jail dataset |
| $\lambda$ | Recovery rate from hospital [1/day] | 0.2<br>Length of hospitalization: *Lognormal*(5,1) | 15 |

The jail provided demographic data about the size of the incarcerated population per day, as well as epidemiological data about confirmed COVID-19 cases over the course of 83 days. We assumed an average reporting delay of six days from first exposure to reported incidence. This accounts for the mean incubation period and a minor delay between symptom onset and COVID-19 test result and isolation. The jail provided data on the age of the infected person, date of positive COVID-19 test, the work or incarceration location of the infected individual, and whether the individual was hospitalized or died as a result of the COVID-19 infection. We used these data to calculate the proportion of symptomatic infections that were hospitalized or died. For each intervention phase, we used the epidemiological data to determine the growth rate, *b*, as the average rate of growth for the entire facility.

We calibrated the transmission rate, *β*, for each intervention phase. We first pseudo-randomly selected values for parameters *ε, α, γ*, and *μ* based on our assumed distributions (Table 1). Then, we calculated *b* for the intervention phase. To find the best-fitting value of *β* for the given parameter set, we implemented an exhaustive search over the range [0,4] in increments of 0.01. We chose the value of *β* which minimized the sum of mean squared error between the reported daily incidence of confirmed COVID-19 cases among incarcerated people and staff in the jail to the daily incidence of symptomatic infected cases in the model for that phase. We calculated the incidence of symptomatic cases using the raw reported incidence before asymptomatic testing. Select asymptomatic testing for incarcerated people began on day 31 and for staff began on day 21. After asymptomatic testing began, we took the minimum of the jail-provided data on the number of symptomatic tests multiplied by the average percentage of positive results of symptomatic tests between days 16-30 (89%) and the raw reported incidence. Based on this estimate, on average, 82% of the reported daily incidence among the incarcerated population was symptomatic after asymptomatic testing began. Because we did not have testing data available for staff, we assumed that 82% of reported new staff cases were symptomatic after on-site testing became available for staff.

We used a simple moving average of the previous five days of incidence to smooth the calibration targets. We assumed that the reported incidence corresponded to the number of incarcerated individuals and staff members who showed symptoms of COVID-19. For each intervention phase, we ran 1,000 Monte Carlo simulations and defined the 95% credible interval of *β* as the range into which 95% of calibrated values of *β* fell.

### Calculation of R_0_ and R_t_

To calculate *R*_0_ and *R*_*t*_, we used the next generation method.^16^ This method utilizes two matrices of partial derivatives of compartments with infected individuals.^17^ In our model, this included exposed, asymptomatic infected, symptomatic infected, quarantined, and hospitalized individuals. The first matrix, *F*, is the rate of appearance of new infections for each compartment. Each element, *f*_*ij*_, of *F* is the partial derivative of any term in which new infections appear in compartment *i* with respect to compartment *j* where *i, j* ∈ [*E, I*_*asym*_, *I*_*sym*_, *Q, H*].

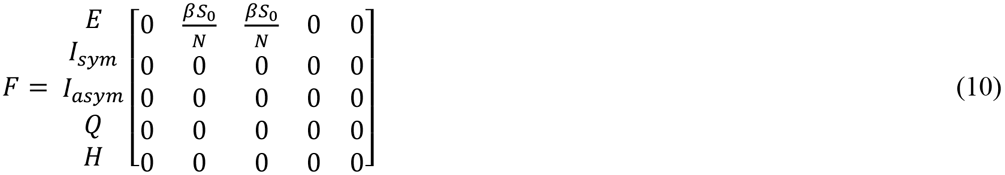

The second matrix, *V*, is the rate of transfer of individuals out of a compartment minus the rate of transfer of individuals into a compartment. Therefore, each element, *v*_*ij*_, of *V* is the partial derivative of the additive inverse of any term other than the appearance of new infections in compartment *i* with respect to compartment *j*. The matrix *V* and its inverse are as follows:

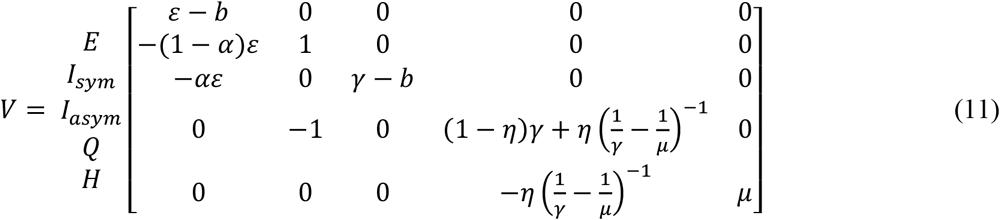

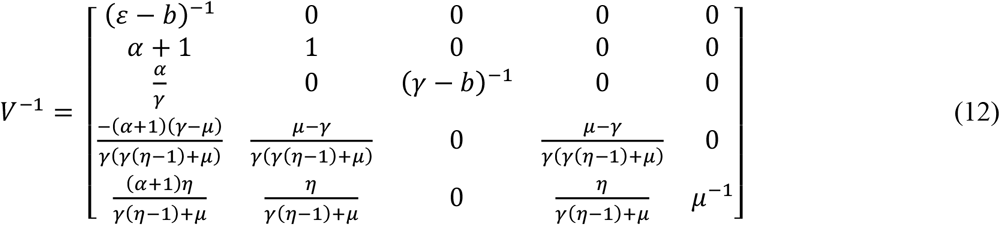

The next generation method calculates *R*_0_ as the dominant eigenvalue of the next generation matrix. The next generation matrix is defined as *FV*^−1^:

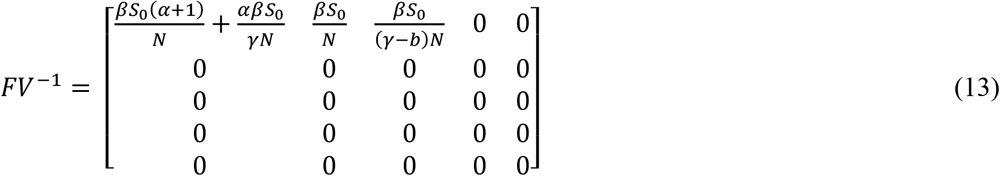

In our model, *FV*^−1^ has only one nonzero eigenvalue, 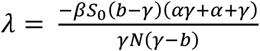. Therefore, *R*_0_ = max (0, *λ*), and since *λ* ≥ 0, *R*_0_ = *λ*. Since *R*_0_ is directly proportional to *β*, we can calculate the values of *R*_0_ of other phases simply by using phase 1 starting conditions combined with the reduced transmission rate.

To find the effective reproduction ratio, *R*_*t*_, at time *t*, we used the next generation method with the same matrices but updated the values of *S* and *β* as appropriate. Because the number of susceptible individuals, *S*, is a function of time, we recalculate *R*_*t*_ each day. The functional form of *R*_*t*_ for our model is as follows:

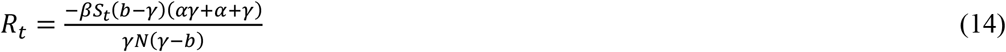

We computed the 95% credible interval of *R*_*t*_ as the range into which 95% of calibrated values of *R*_*t*_ fell.

This study was deemed exempt from IRB review by the Yale Human Investigation Committee as we received completely anonymized data from the jail.

## RESULTS

Daily reported incidence of COVID-19 in the jail was highly variable, ranging from 0 to 67. The mean absolute error of the model compared to the simple moving average was 19% (Figure 3).

**Figure 3.**
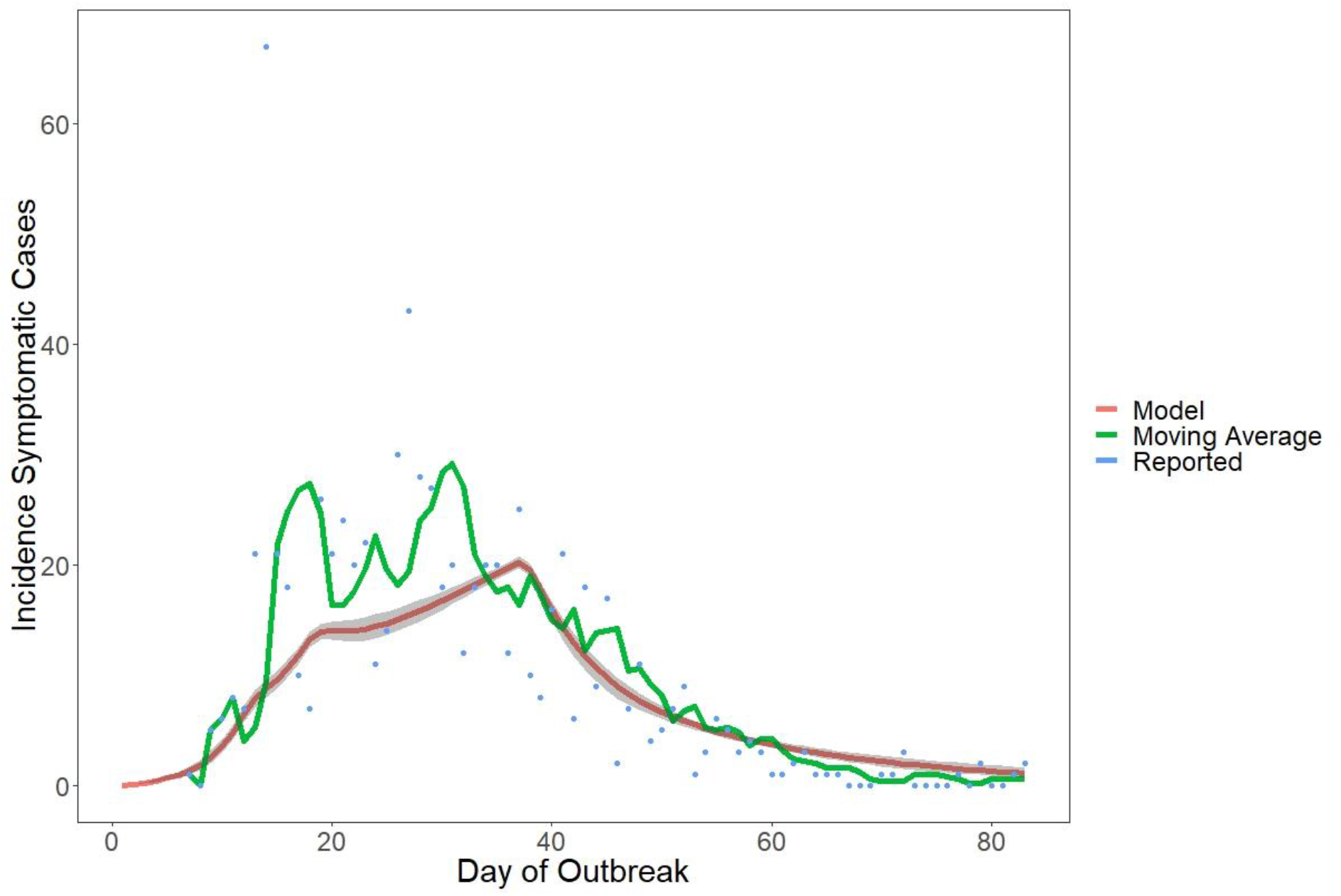
Comparison of the incidence of symptomatic cases in the model with reported COVID-19 incidence at the jail.

### Transmission Rates

In following the initial CDC recommendations for correctional facilities (phase 1), the baseline transmission rate (*β*) was 1.89 (95% Credible Interval (CrI): 1.44-2.44) (Figure 4). After depopulation began (phase 2), the transmission rate was *β*= 0.83 (95% CrI: 0.66-1.06). This represents a 56% decrease in the transmission rate from phase 1. After the increase in single-occupancy cells (phase 3), the transmission rate was *β*= 0.41 (95% CrI: 0.30-0.56), a 51% decrease from phase 2. Finally, the transmission rate after testing of asymptomatic individuals began (phase 4) was *β*= 0.11 (95% CrI: 0.06-0.20), a 73% decrease from phase 3. All of these reductions are statistically significant.

**Figure 4.**
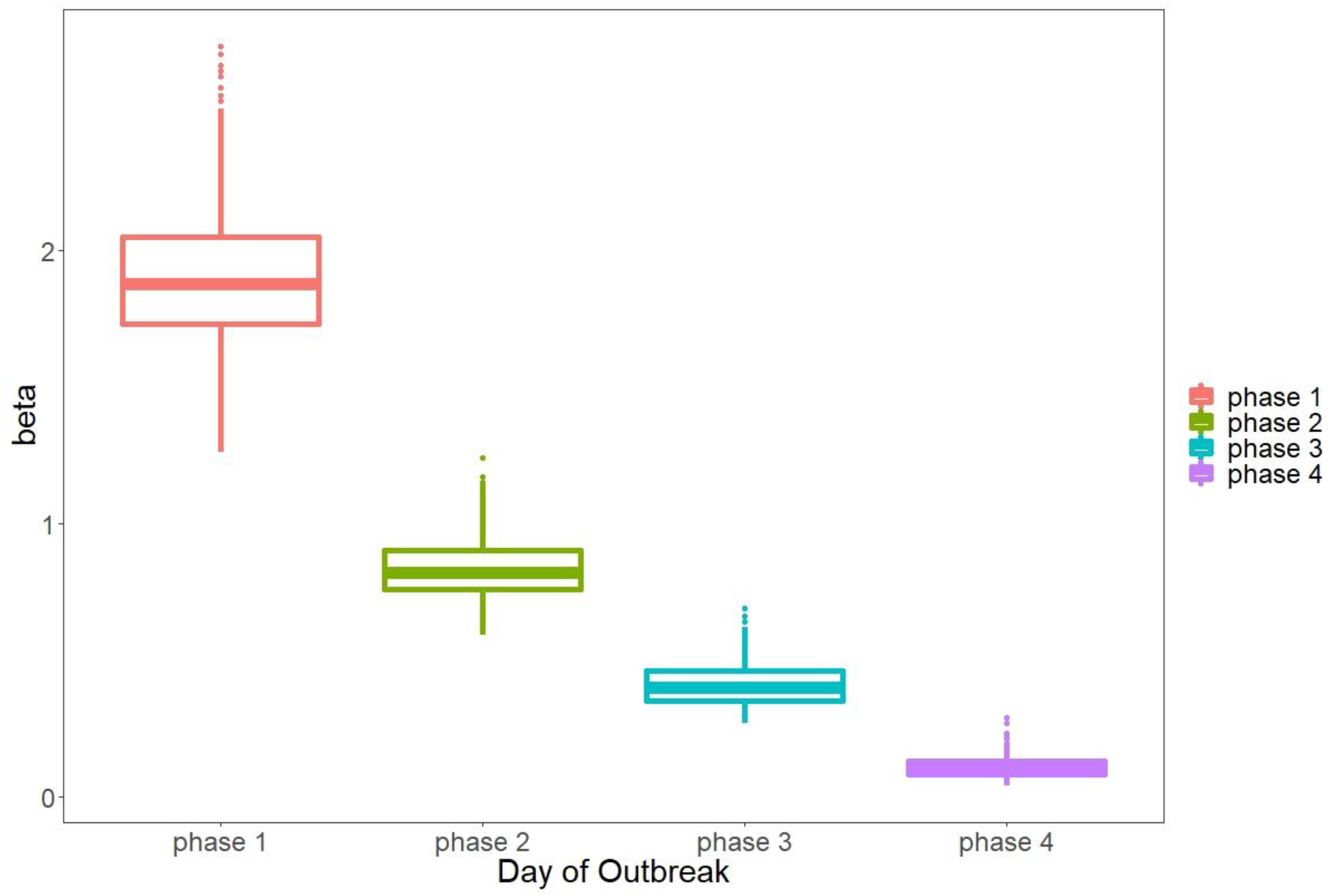
Calibrated values of the transmission rate *β* for different outbreak phases (Phase 1: initial outbreak, Phase 2: depopulation, Phase 3: increased single celling, Phase 4: widespread testing of asymptomatic incarcerated individuals).

### Reproduction Ratios

The estimated value of *R*_0_ was highest in phase 1, during the first 11 days of the outbreak (Table 2). For this phase, we estimate *R*_0_ = 8.23 (95% CrI: 5.01-12.90) (Table 2). We estimate *R*_0_ of each phase in a completely susceptible population as if the outbreak had begun with the values for *β* which correspond to each phase: *R*_0,*phase* 2_ = 3.58 (95% CrI: 2.46-5.08), *R*_0,*phase* 3_ = 1.72 (95% CrI: 1.41-2.12), and *R*_0,*phase* 4_ = 0.45 (95% CrI: 0.32-0.59). The effective reproduction ratio, *R*_*t*_, decreased over time as the susceptible population shrank, the transmission rate changed, and different interventions were implemented (Figure 5). For the entire jail, we estimate that the interventions may have reduced the effective reproduction ratio *R*_*t*_ below 1 about five weeks after the outbreak began (on day 37).

**Table 2.** Intervention Effects: Estimated Transmission Rates (*β*), Effective Reproduction Ratios (*R*_0_), and Disease Cases for each Outbreak Phase.

| Phase | Time Range in Days | $\beta$<br>(95% CrI) | $R_0$<br>(95% CrI) | Reduction in $\beta$ and $R_0$ from Previous Phase | Expected Total Symptomatic Cases, Day 83*<br>(95% CrI) | Expected Total Hospitalizations, Day 83*<br>(95% CrI) | Expected Total Deaths, Day 83*<br>(95% CrI) | Expected Total Cases, Day 200*<br>(95% CrI) |
| --- | --- | --- | --- | --- | --- | --- | --- | --- |
| 1: Initial outbreak | 1 – 11 | 1.89<br>(1.44 - 2.44) | 8.25<br>(5.01 - 12.90) |  | 3,867<br>(2,742 - 5,044) | 541<br>(384 - 706) | 38<br>(29 - 47) | 6,372<br>(6,318 - 6,437) |
| 2: Depopulation | 12 – 17 | 0.83<br>(0.66 - 1.06) | 3.58<br>(2.46 - 5.08) | 56% | 2,520<br>(1,940 - 3,088) | 353<br>(272 - 432) | 24<br>(20 - 28) | 4,055<br>(3,666 - 4,294) |
| 3: Increased single celling | 18 – 36 | 0.41<br>(0.30 - 0.56) | 1.72<br>(1.41 - 2.12) | 51% | 1,447<br>(1,224 - 1,654) | 203<br>(171 - 232) | 12<br>(11 - 13) | 2,950<br>(2,331 - 3,521) |
| 4: Widespread testing of asymptomatic incarcerated individuals | 37 – 83 | 0.11<br>(0.06 - 0.20) | 0.45<br>(0.32 - 0.59) | 73% | 642<br>(592 - 692) | 90<br>(83 - 97) | 3.9<br>(3.6 - 4.1) | 1,121<br>(904 - 1,433) |

\* Assuming the value of $\beta$ estimated for this intervention phase occurs during all subsequent days.

**Figure 5.**
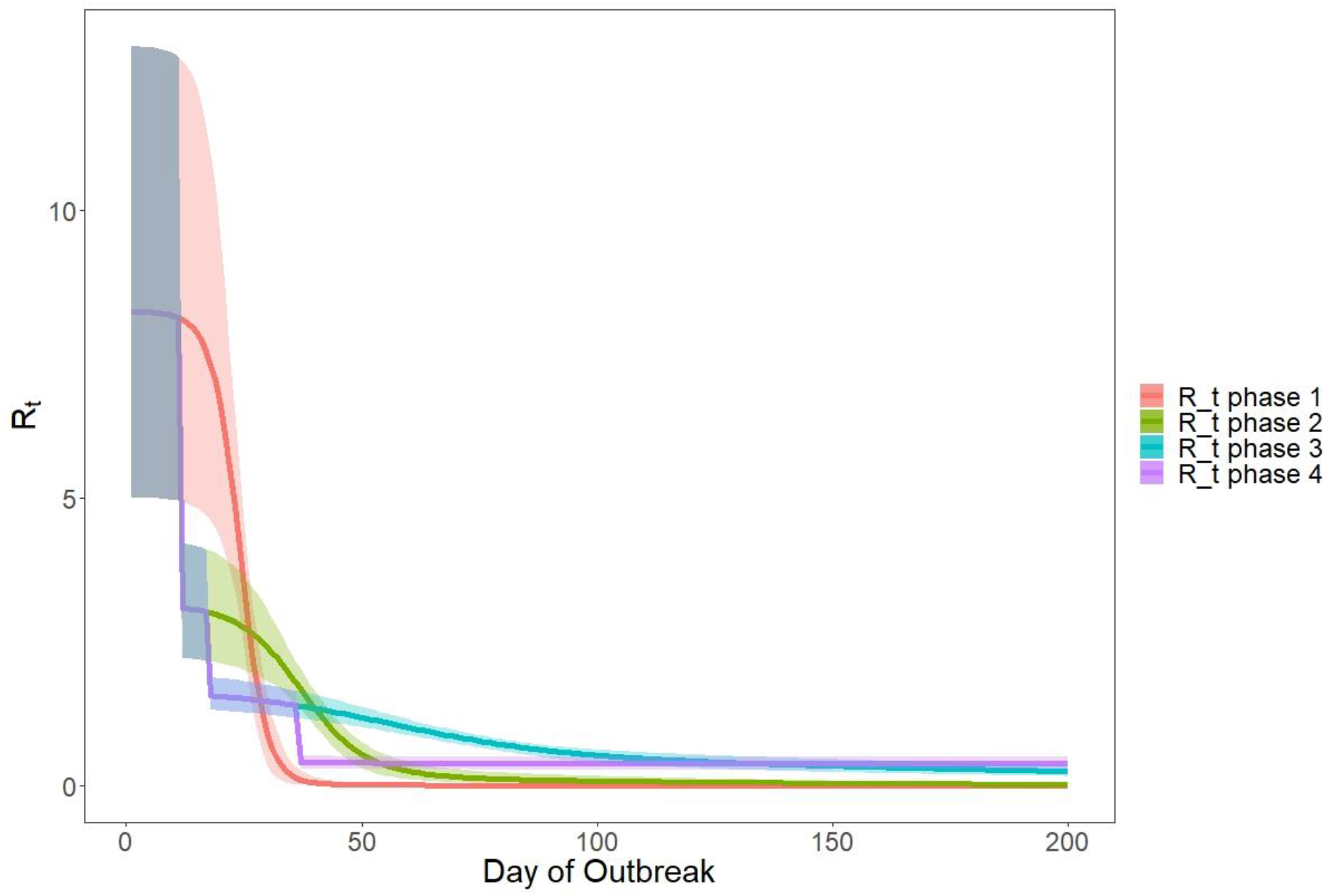
Calculated values of the effective reproduction ratio *R*_*t*_ for all intervention phases (Phase 1: initial outbreak, Phase 2: depopulation began, Phase 3: increased single celling, Phase 4: widespread testing of asymptomatic incarcerated individuals).

### Averted Infections, Hospitalizations, and Deaths

Table 2 shows the expected total symptomatic cases on day 83 and expected total cases on day 200, assuming that the estimated transmission rate for a particular outbreak phase holds over all subsequent days. Over the first 83 days of the outbreak, the jail reported 778 symptomatic cases among incarcerated individuals and staff. Our model predicts 642 symptomatic cases (95% CrI: 592-692), 90 hospitalizations (95% CrI: 83-97), and 4 deaths (95% CrI: 3.6-4.1) over this same time period (Figure 6). Our estimate is 17% less than the number of reported cases that were symptomatic. Compared to what could have happened with only CDC recommended interventions, the model predicts a reduction of over 3,200 symptomatic cases, 450 hospitalizations, and 30 deaths over 83 days. This suggests that the combination of interventions (depopulation, increased single celling, and large-scale asymptomatic testing of incarcerated individuals) in addition to standard CDC COVID-19 mitigation strategies led to an 83% reduction in predicted symptomatic cases and hospitalizations and an 89% reduction in predicted deaths.

**Figure 6.**
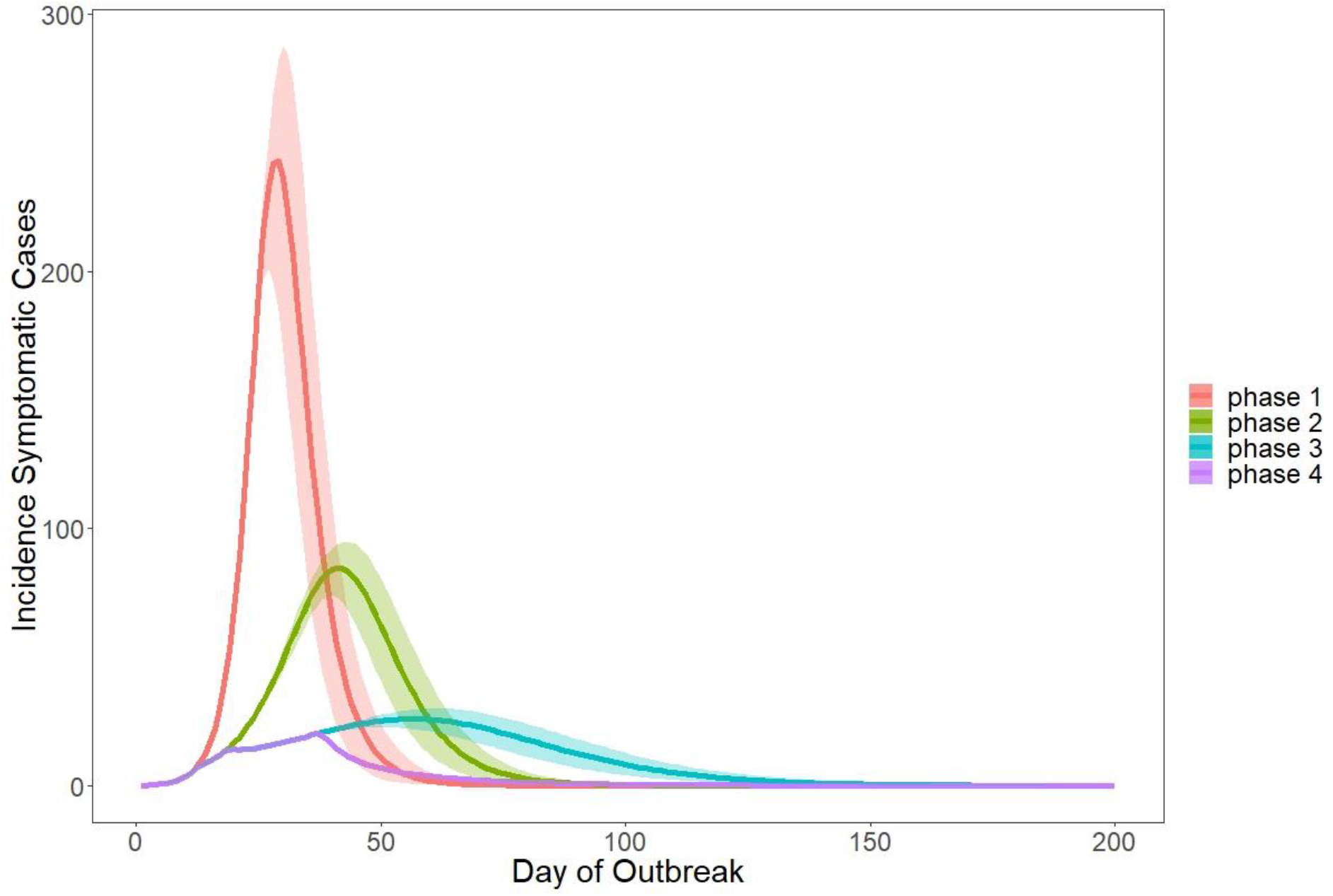
Projected incidence of symptomatic cases for all intervention phases (Phase 1: initial outbreak, Phase 2: depopulation began, Phase 3: increased single celling, Phase 4: widespread testing of asymptomatic incarcerated individuals).

## DISCUSSION

Using a stochastic compartmental model, we estimate that depopulation efforts, single celling and asymptomatic testing are important interventions, in addition to those recommended by the CDC to reduce COVID-19 transmission in jails. We estimate that the actions taken by the jail reduced potential new cases by approximately 83% over 83 days, and this may have averted over 450 hospitalization and 30 deaths among those who work and live in jails.

Given these findings, depopulation efforts should be a primary strategy for COVID-19 mitigation in jails. Reductions in detained populations to prevent disease transmission is best achieved by both decreasing the number of new intakes and increasing the number of releases. This requires that authorities that control jail admissions (including police departments, judges, and in some cases correctional departments) and jail releases (including judges, lawyers and community bail funds) both focus on promoting depopulation efforts to mitigate COVID-19 transmission.

Our data also suggest that jails should focus on single celling to mitigate COVID-19. To be clear, single celling does not imply solitary confinement but rather placing one person in a 6 × 9-foot cell to increase physical distancing in correctional facilities.^18^ Given physical crowding in many facilities even when overall incarcerated populations are at record lows, increasing access to single-occupancy cells will not be feasible without depopulation efforts, and as supported by our model, will not lead to a contained transmission rate alone. Facilities unable to appropriately place individuals in single cell without relying on solitary confinement should consider depopulation as a preferred strategy. Implementing all of these measures will require interagency coordination to achieve the full public health impact. Further, by enacting these measures, correctional facilities may contribute to managing transmission in the surrounding community as well, as several recent studies have documented jails as drivers of community spread of COVID-19.^19,20^

Lastly, asymptomatic testing is an important component to COVID-19 mitigation strategies. In this jail, they focused on asymptomatic testing through contact tracing of people who tested positive, but much more research needs to be conducted on who should be tested and under what circumstances, including whether mass testing is effective, when individuals should be tested (upon entry, upon release, only for contact tracing, or in regular intervals), and whether certain community rates should guide whether asymptomatic people warrant testing in corrections. While widespread asymptomatic testing may not be indicated in a jail without community cases, when community cases are present, asymptomatic testing should be strongly recommended.

National and international health agencies, such as the CDC and the World Health Organization, should address depopulation, single celling, and asymptomatic testing in future guidance for detention facilities and how best to implement these measures. Correctional facility administrators will need to also consider how to best mitigate the challenges that come with any of these strategies. For example, coordination of health care and social services organizations prior to release should be prioritized and considerations of testing when releasing individuals as part of depopulation efforts.

Our analysis has several limitations. We used a compartmental model which assumes homogeneous mixing among the entire population. Correctional facilities in reality do not exhibit homogeneous mixing, especially across divisions. Our model does not have the granularity to capture the influence of individuals on transmission dynamics. Our model assumes a relatively stationary population and only accounts for mixing within the jail. In reality, jail populations are highly variable with frequent intakes and releases. Jailed individuals also have variable daily routines, such as where they eat or exercise, which are not accounted for in our model. We did not account for possible false positives, misdiagnosis, overreporting, or underreporting in the dataset. Finally, the many interventions undertaken by the jail make it difficult to determine the causal influence of any one particular intervention.

Importantly, these limitations influence our estimates of *β* and *R*_0_. We model the jail as a closed system and thus neglect exogeneous infection (e.g., staff or new intake incarcerated individuals who contracted the disease in the community) that likely entered the jail before large-scale testing efforts. Because our analysis assumed that all new infections arise from internal transmission, we likely overestimate the true values of *β* and *R*_0_, particularly in the early phases of the epidemic in the jail. Thus, conclusions resulting from our analysis should focus on the relative reductions of *β* and *R*_0_ rather than the precise estimates of these values.

Despite the limitations of our analysis, we conclude that it is possible to mitigate the spread of COVID-19 even in correctional settings, where standard social distancing practices are difficult to achieve, by implementing depopulation strategies, promoting increased single celling, and asymptomatic testing with appropriate isolation. The large estimated reduction in the transmission rate (≥ 50%) from these three intervention strategies is comparable to standard social distancing measures in a community setting.^21^ As states and the federal government are focused on re-opening economies, strategies should be devised to protect those who are incarcerated and those who work in corrections by further limiting population increases so that future outbreaks are averted.

## Data Availability

The data was made available through the cooperation of an anonymous large urban jail under a data sharing agreement. Please contact GM for questions regarding sharing availability.

## Acknowledgments

We acknowledge the anonymous jail for their prompt collaboration and transparency in providing detailed data on COVID-19 transmission. At the time that this work was conducted, G.M. was supported by grant number T32HS026128 from the Agency for Healthcare Research and Quality and L.P. was partially supported by the Veterans Health Administration. The content is solely the responsibility of the authors and does not necessarily represent the policy or views of the Agency of Healthcare Research and Quality, the Veterans Health Administration or the United States Government.

